# Cardiorespiratory exercise and self-management early after stroke to increase daily physical activity: results from a stepped-wedge cluster randomized trial

**DOI:** 10.1101/2024.04.24.24306073

**Authors:** Augustine J. Devasahayam, Ada Tang, Denise Taylor, Elizabeth L. Inness, Rebecca Fleck, Esmé French, David Jagroop, Cynthia Danells, Avril Mansfield

## Abstract

**Trial design:** Stepped-wedge cluster superiority randomized controlled trial.

**Objective:** This study aimed to determine if Promoting Optimal Physical Exercise for Life (PROPEL) program increases participation in physical activity up to six months post-discharge from stroke rehabilitation, compared to participation in group cardiorespiratory exercise (GCE) alone.

**Methods:** People with sub-acute stroke participated in the PROPEL (n=107) or GCE (n=65) intervention phases. The primary outcome was adherence to physical activity guidelines over seven days at six months post-discharge from rehabilitation. Secondary outcomes were exercise self-efficacy (Short Self-Efficacy for Exercise scale), exercise-related beliefs and attitudes (Short Outcome Expectation for Exercise scale), and perceived barriers to physical activity (Barriers to Being Active Quiz).

**Results:** Fifty seven participants (PROPEL, n=29; GCE, n=28) were included in the analysis. At six months post-discharge, 6/17 PROPEL participants and 9/22 GCE participants met the guidelines for intensity and duration of physical activity; the odds of meeting the physical activity guidelines did not differ between phases (p>0.84). PROPEL participants reported higher self-efficacy for exercise than GCE participants (p=0.0047).

**Conclusions:** Participation in the PROPEL increases self-efficacy for exercise compared to GCE alone after stroke. However, increased self-efficacy for exercise did not increase the odds of meeting physical activity guidelines.

**Trial registration:** NCT02951338

## INTRODUCTION

People who have had a stroke often have low cardiorespiratory capacity (1), which can make activities of daily living more effortful and negatively affect quality of life (2). Regular cardiorespiratory exercise can increase functional capacity, improve the ability to carry out daily activities, and improve quality of life post-stroke (3). Physically active people who have had a stroke report higher satisfaction with life than those who are less active (4). Cardiorespiratory exercise is recommended to reduce the risk of another stroke and/or other cardiovascular events (3). Even in the early stages after a stroke, cardiorespiratory exercise is beneficial (5) and can be incorporated into routine stroke rehabilitation (6). Cardiorespiratory exercise may be implemented as part of inpatient stroke rehabilitation, but due to short lengths of stay (often less than 30 days (7)), the duration of exercise is likely too short to bring about meaningful changes in cardiorespiratory fitness (8). Therefore, people with stroke are encouraged to participate in longer-term cardiorespiratory exercise after discharge to improve and maintain cardiorespiratory fitness (9).

Unfortunately, most people with stroke are physically inactive following discharge from rehabilitation. People with stroke discontinue more than half of the physical activities they had engaged in before the stroke (4). People with stroke living in the community walk, on average, approximately 4300 steps per day (10), which is much lower than the 6000-6500 steps per day recommended for people who are living with a disability and/or chronic illness (11). Conversely, people with stroke who take more than 6025 steps per day have reduced risk of secondary vascular events post-stroke (12).

Given the low levels of physical activity among people with stroke, they may benefit from referral to supervised exercise programs, such as adapted cardiac rehabilitation, after inpatient stroke rehabilitation (13-15). However, access to, enrollment, and attendance in these programs are often poor. For example, only 42% of people who were referred to cardiac rehabilitation after stroke enrolled (16) and 38% of those who enrolled in cardiac rehabilitation attended less than half of the exercise sessions (17). Therefore, interventions to promote longer-term participation in physical activity and planned exercise post-stroke are necessary.

Interventions aimed at increasing physical activity after a stroke, particularly for people who have completed inpatient rehabilitation, often incorporate behavior change principles (18). There is an opportunity to offer cardiorespiratory exercise along with strategies for modifying behaviour post-stroke to not only increase cardiorespiratory fitness but also to improve long-term physical activity participation after returning to the community (19). We developed Promoting Optimal Physical Exercise for Life (PROPEL), a combined cardiorespiratory and behaviour change program, to help people after stroke have the knowledge, skills, and self-efficacy to maintain exercise and physical activity after discharge from rehabilitation (20, 21).

The primary aim of this study was to determine the effect of PROPEL, delivered during stroke rehabilitation, on participation in self-directed exercise and physical activity after discharge from rehabilitation. The secondary aims were to determine the effect of PROPEL on self-efficacy and outcome expectations for exercise and barriers to participating in exercise. We hypothesized that more people with stroke who complete PROPEL will meet the recommended intensity and duration of self-directed physical activity in the community six months post-discharge from rehabilitation (22), compared to those who complete group cardiorespiratory exercise (GCE) only. The secondary hypotheses were that people with stroke who complete PROPEL will report higher self-efficacy and outcome expectations for exercise, and fewer reported barriers to exercise participation.

## METHODS

### Trial design

This was a multi-site, prospective, assessor-blinded, continuous recruitment, stepped-wedge cluster superiority randomized controlled clinical trial (21). Sites conducted the control intervention (GCE) and the experimental intervention (PROPEL) as shown in the adapted CONSORT Stepped Wedge Cluster Randomized Controlled Trial Flow Chart (Figure 1). The following changes were made to the protocol (21) after study initiation: the number of sites was reduced (from six to five); the duration of the intervention phases at each site was modified; eligibility criteria were changed; and how outcomes were defined were modified. Explanations for these changes are provided in the relevant sections below. Throughout the study period, potential participants were continuously screened and assigned to either the GCE or PROPEL intervention based on the program being administered at the time of their enrolment into the study intervention. Participants were enrolled into the study at the end of the GCE or PROPEL program and provided written informed consent for study participation. The trial is reported according to the CONSORT extension for stepped wedge cluster randomized trials (23), and the CONSERVE-CONSORT checklist for trials modified due to the coronavirus disease pandemic (COVID-19) and other extenuating circumstances (24). Modifications to the study methods after the onset of COVID-19 were implemented by the principal investigator, in consultation with the study team, and were approved by the research ethics boards of the respective study sites.

**Figure 1:**
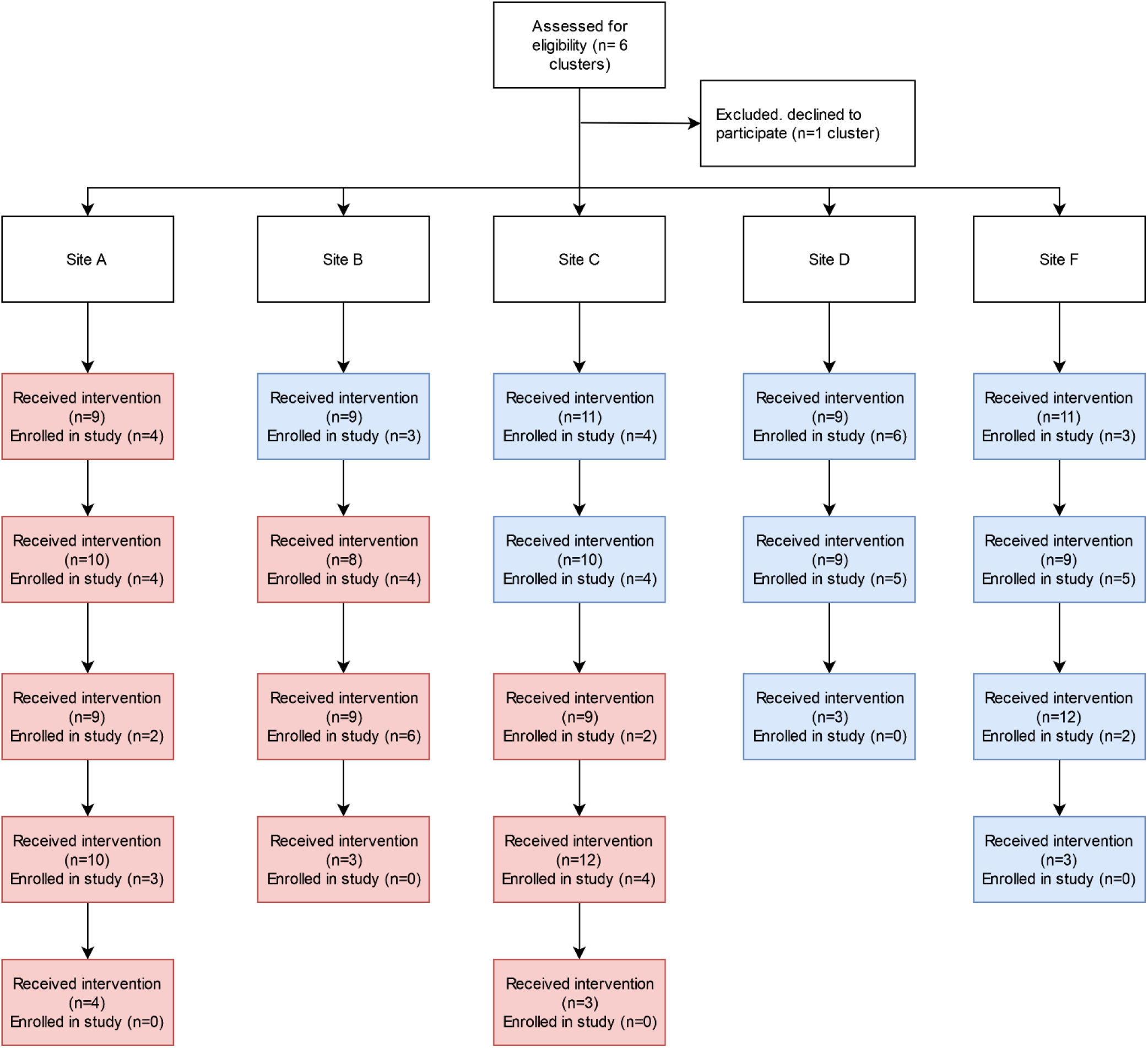
CONSORT stepped wedge cluster randomized controlled trial flow chart. The red boxes indicate the PROPEL phase and the blue boxes indicate the GCE phase. Each box represents up to 3 intervention groups.

### Participants

#### Eligibility criteria

To qualify for referral to GCE or PROPEL, patients must have been admitted to one of the rehabilitation hospitals, either an inpatient or outpatient program, following a stroke and have had sufficient cognitive capacity to understand and follow instructions and to communicate any adverse symptoms (such as pain or excessive exertion) during exercise. Exclusion criteria were: (a) medical conditions that limited their ability to exercise, such as uncontrolled hypertension, uncontrolled diabetes, cardiovascular co-morbidities that limited exercise tolerance (e.g., heart failure, abnormal blood pressure responses or ST-segment depression > 2 mm, symptomatic aortic stenosis, complex arrhythmias), unstable angina, orthostatic hypotension that exceeded 20 mm Hg, or musculoskeletal pain or impairments; and/or (b) cardiovascular abnormalities detected during the submaximal exercise test. Patients who completed either GCE or PROPEL as part of their routine care at one of the five rehabilitation hospitals were eligible for participation in the study. Patients were not invited to participate in the study if they had cognitive impairments that prevented participation in unsupervised exercise. Initially, we excluded patients who attended less than 50% of GCE or PROPEL training sessions and/or attended less than four of the six group discussion sessions (for the PROPEL phase); however, these exclusion criteria were removed in August 2019 due to low rates of referral to GCE/PROPEL and consequently low enrolment into the study.

#### Study setting

Participants were recruited from one of five rehabilitation hospitals in Ontario: Hamilton Health Sciences, Hamilton; Sunnybrook Health Sciences Centre – St. John’s Rehab, Toronto; St. Joseph’s Care Group, Thunder Bay; Toronto Rehabilitation Institute – University Centre, Toronto; and Toronto Rehabilitation Institute – Rumsey Centre, Toronto. One additional site had been recruited for the study, but this site withdrew as they were unable to hire personnel to support the study. This study was approved by the research ethics boards of: (a) Hamilton Health Sciences, Hamilton, Ontario (approval number: 2274); (b) Sunnybrook Health Sciences Centre, Toronto, Ontario (approval number: 472-2016); (c) St. Joseph’s Care Group, Thunder Bay, Ontario (approval number: 2016011); (d) University Health Network, Toronto, Ontario (approval number: 16-5916); (e) West Park Healthcare Centre, Toronto, Ontario (approval number not provided in the letter); (f) Thunder Bay Regional Health Sciences Centre, Thunder Bay, Ontario (approval number: 2016139). Each site was staffed by a research assistant (RA) and a research physiotherapist (PT). The RA was responsible for recruiting participants and collecting data, while the PT administered the interventions. PTs at each site monitored participants for adverse events during the intervention.

### Interventions

Depending on the study phase, patients were referred to either GCE or PROPEL by their primary treating physiotherapist. Interventions were delivered by the research PT in a group format, with at least three patients per group. Patients could be referred to the intervention groups during inpatient or outpatient rehabilitation.

### Control intervention: group cardiorespiratory exercise (GCE)

The GCE phase involved individually prescribed cardiorespiratory exercise (based on sub-maximal or maximal cardiorespiratory capacity tests) 3 days per week for 6 weeks, using modalities such as a recumbent stepper, cycle ergometer, or treadmill (21). In general, each exercise session included 3 to 5 minutes of low-intensity exercise to warm up, 20-30 minutes of exercise at an intensity of 50% to 70% of the age-predicted maximum heart rate or a rating of 3/10 (moderate) on the Borg category ratio (CR-10) scale (25), and 3-5 minutes of low-intensity exercise to cool down. However, the specific exercise prescription could deviate from these general principles, depending on factors such as patient tolerance for exercise, comorbid conditions, and patient preferences.

### Experimental intervention: Promoting Optimal Physical Exercise for Life (PROPEL)

The PROPEL phase involved individually prescribed cardiorespiratory exercise sessions 3 days per week for 6 weeks, as described for GCE (21). In addition, participants in PROPEL participated in one-hour group discussion sessions once per week, focusing on acquiring self-management skills for exercise in preparation for discharge from rehabilitation. The specific objectives of these discussion sessions during PROPEL were identifying and solving barriers to participating in exercise, understanding the general and personal benefits of participating in exercise, and finding realistic and personalized strategies for adding exercise into daily life routine (21).

### Outcomes

#### Primary outcome measure: Physical activity

Physical activity over seven consecutive days was assessed at three time points: (a) 1 month, (b) 4 months, and (c) 6 months post-discharge from rehabilitation. Participants wore a commercial wrist-worn step counter and heart rate monitor (Fitbit Charge HR, Fitbit Inc., San Francisco, USA) for seven consecutive days at each time point. Participants who used a standard or a wheeled walker for ambulation also wore an activity monitor on the ankle (Fitbit One; 26). Heart rate and step count data were not displayed on the Fitbit devices so that participants did not receive real-time feedback about their physical activity. At the end of the 7-day physical activity monitoring period, the blinded RA completed the 13-item Physical Activity Scale for Individuals with Physical Disabilities (PASIPD) questionnaire over the telephone (27). The total PASIPD scores were calculated by multiplying the metabolic equivalent (MET) value for each activity by the average number of hours per day spent on each activity and then adding the scores from all activities (in total MET hours/day) (27, 28).

Data from the activity monitor (step count and heart rate) and PASIPD questionnaire were used to determine if participants met the recommended intensity and duration of physical activity in the community; that is, at least 150 minutes per week of moderate to vigorous intensity exercise (29), or at least 75 minutes per week of vigorous-intensity exercise (22), or took at least 6000 steps per day (11, 12, 30).

The criterion to meet the physical activity guidelines based on step count was modified from the protocol (21). We originally intended to use the ‘active minutes’ output by Fitbit to determine if participants met the guidelines for intensity and duration of physical activity in the community based on step data. However, Fitbit often output 0 minutes of activity, even when step counts and heart rate data indicated that participants were active. Therefore, we used a threshold of 6000 steps from the step counter as a minimum of 6000-6500 steps/day is recommended for people with disabilities (11, 30) and is associated with a reduced risk of recurrent cardiovascular events (12).

Participants were deemed to meet the physical activity guidelines based on heart rate data if their heart rate was within a moderate-to-vigorous intensity range (>40% heart rate reserve) for at least 150 minutes/week, or within a vigorous intensity range (>60% heart rate reserve) for at least 75 minutes/week (22). This criterion was also modified from the protocol (21). We initially intended to determine if participants met the guidelines for physical activity based on age-predicted maximum heart rate. However, we noted that extremely low age-predicted maximum heart rates for participants using beta-blockers (calculated using an adjusted formula for beta-blockers (31)) led to participants being within the moderate and/or vigorous intensity heart rate range for almost the entire day.

Alternatively, heart rate reserve, which considers both maximum heart rate and resting heart rate, resulted in more realistic times spent within the target range for our participants. Resting heart rate was obtained from the Fitbit data during the recording week. If participants completed a maximal cardiopulmonary exercise test during rehabilitation and were confirmed to reach their physiological maximum on this test, their peak heart rate from the cardiopulmonary exercise test was used as the maximum heart rate. Alternatively, the maximum heart rate was either the age-predicted maximum heart rate (32, 33) or the highest heart rate recorded by Fitbit, whichever was higher.

Participants were deemed to meet the physical activity guidelines based on the PASIPD if they reported at least 150 minutes of moderate-to-vigorous intensity exercise (PASIPD items related to moderate sport and recreational activities, strenuous sport and recreational activities, and exercise to increase muscle strength and endurance) or at least 75 minutes of vigorous-intensity exercise (PASIPD item related to strenuous sport and recreational activities).

#### Secondary outcomes

Exercise self-efficacy was evaluated using the Short Self-Efficacy for Exercise (SSEE) scale (34), which is a four-item questionnaire where participants were asked to rate their confidence on a five-point scale when exercising through pain, fatigue, being alone, and feeling depressed. Beliefs and attitudes related to exercise were evaluated using the Short Outcome Expectation for Exercise (SOEE) scale (34) which is a five-item questionnaire where participants were asked to rate their beliefs and attitudes towards exercise on a five-point scale. The SSEE and SOEE were obtained upon study enrolment (i.e., immediately after completing GCE or PROPEL). Perceived barriers to physical activity were assessed at the one month post-discharge time point using the Barriers to Being Active Quiz (BBAQ; (35), which consisted of a 21-item questionnaire, where participants indicated their level of agreement with statements related to barriers to exercise, split into 7 categories of barriers: lack of time, social influence, lack of energy, lack of willpower, fear of injury, lack of skill, and lack of resources, with each item scored on a scale of 0 to 3. The total score for each category was the sum of the scores for all items within that category (i.e., maximum category score of 9). The number of significant barriers was the number of categories with a score of 5 or higher (35).

#### Participant characteristics

The following participant characteristics were obtained through chart review or directly from the participant: age, sex, time since stroke (calculated at the time of the start of the GCE/PROPEL intervention), lesion location, mobility status, and medical history. At enrollment, the National Institutes of Health Stroke Scale (NIH-SS; 36), Chedoke-McMaster Stroke Assessment (CMSA) leg and foot scores (37), and Montreal Cognitive Assessment (MoCA; 38) were obtained by the RA or PT. If these measures were recently conducted as part of clinical care (within 1 week of study enrolment), the scores were extracted from the hospital charts. The Schmidt retrospective physical activity scale was used to evaluate premorbid exercise behaviour, where participants indicated their average amount of time (hours/day) before stroke spent in sedentary activities (such as watching television) and in physical activities or exercise (39).

### Sample size

We expected that approximately 25% of patients who completed GCE (40) and 50% of those who completed PROPEL (20) would be classified as “active”. A sample size of 96 participants per phase (GCE and PROPEL) was determined to provide 80% power to detect a 25%-50% difference at an alpha level of 0.05 for six study sites, considering an intra-cluster correlation of 0.05 (41). The goal was to enroll a total of 120 participants per phase, accounting for a 20% dropout rate.

### Randomization

#### Sequence generation, allocation concealment, and implementation

The time at which each site transitioned from GCE to PROPEL was determined by drawing site names at random. Intervention allocation was performed by the principal investigator, who was not directly involved in study recruitment or data collection, at the start of the study. The site leads and PTs at each study site who enrolled participants into clusters were informed of the transition from GCE to PROPEL approximately 3 months before the transition to allow for sufficient time for training and planning.

#### Blinding

Participants cannot be blinded to intervention allocation, although they were not aware of the existence of another intervention arm. Assessors (RA at each site) who collected data, including administering questionnaires, were unaware of the time at which the site transitioned from GCE to PROPEL. While it is more likely that a given site would be allocated to GCE at the start of the study period, and to PROPEL at the end of the study, the inclusion of two sites that always completed either GCE or PROPEL created uncertainty in intervention allocation at all time points. Using objective methods to collect data pertaining to the primary outcome (i.e., heart rate and activity monitor) post-discharge from rehabilitation helped to protect against bias if assessors inadvertently became unblinded. Furthermore, participants were blinded to their heart rate and step count by turning off the display feature on the data collection device during the activity log period of the evaluation.

### Statistical methods

Missing physical activity data were imputed for participants still enrolled in the study at each time point (42). Missing data were not imputed for participants who had withdrawn from the study at each time point as missing data due to withdrawal were likely missing not at random. Site (i.e., cluster), cohort descriptors and baseline data (i.e., age, sex, pre-morbid physical activity CMSA scores, NIH-SS scores, BBAQ scores, SSEE scores, FAC scores), and non-missing physical activity data were used to impute missing data. Multiple logistic regression, with fixed effects of phase and time and a random effect of site was used to test the primary hypothesis (i.e., odds of meeting guidelines for physical activity). Multiple linear regression was used to compare PASIPD scores, heart rate data (time in moderate-vigorous, and time in vigorous heart rate ranges), and step counts between phases, with site and time included in the models. To test the secondary hypotheses, the Wilcoxon Rank-Sum (Wilcoxon Mann-Whitney U) test was used to compare SSEE, SOEE, and BBAQ scores between the groups.

## RESULTS

### Participant flow and recruitment

The study ran from March 2017 to March 2020. As noted above, one site (Site E) withdrew from the study without running any study interventions. Four sites initiated the intervention between March and July 2017. Site D delayed starting the intervention until August 2018 as another exercise study was taking place at that site, and it was believed that the current study interventions would conflict with that other study. The sites ended the study interventions in March 2020 due to the onset of the COVID-19 pandemic. While the sites attempted to continue the study after pandemic control measures were eased, they were ultimately unable to restart the intervention groups. Across all sites, 172 patients were referred to the exercise groups. Of these, 59 consented to participate in the study. Two participants withdrew from the study before completing any data collection and were not included in the analysis. The participant recruitment flow chart as per study phases is shown in Figure 2. Participant baseline characteristics are shown in Table 1.

**Table 1:**
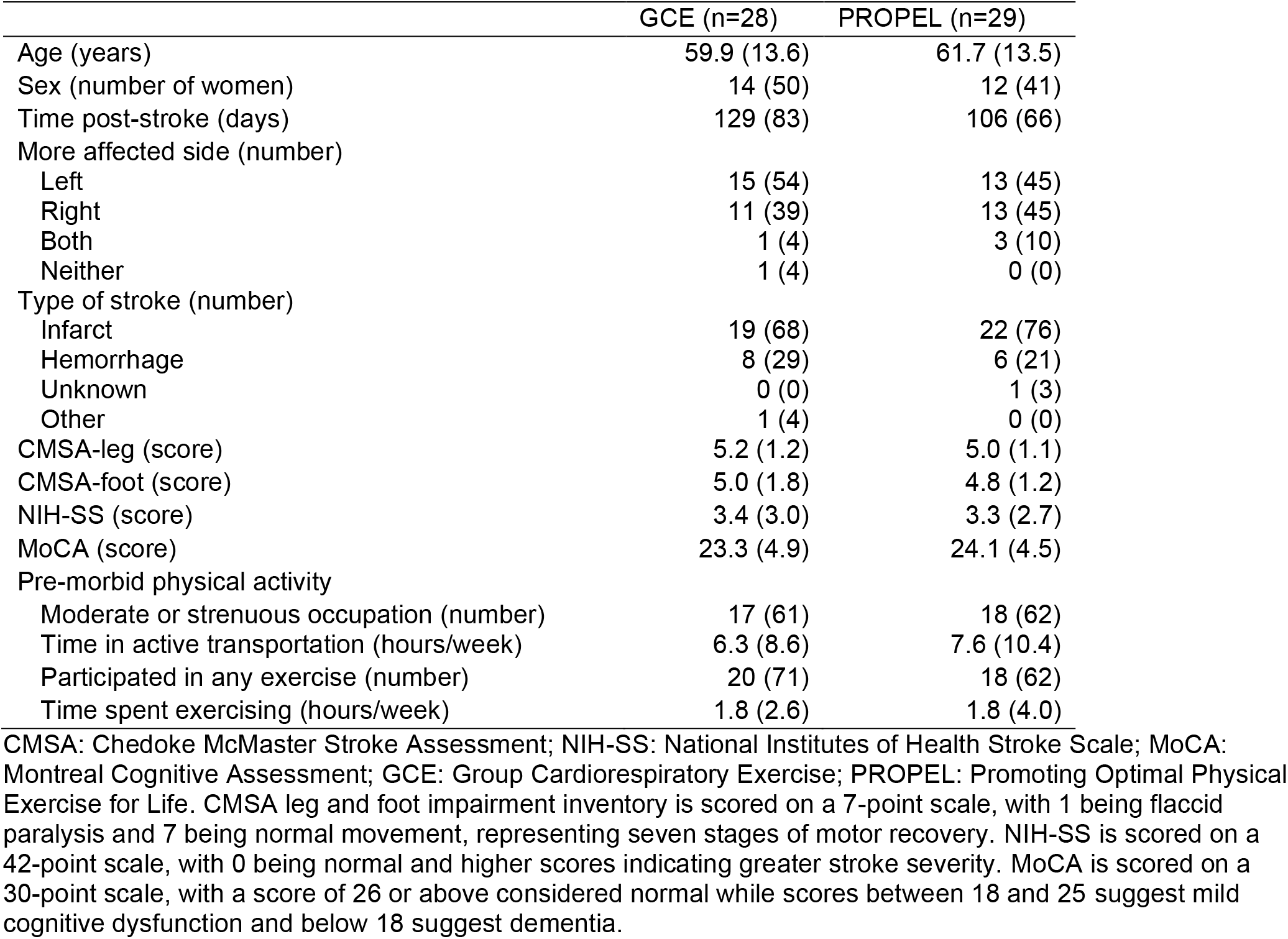
Baseline characteristics. Values presented are means with standard deviations in parentheses for continuous variables, or counts with percentages in parentheses for categorical variables.

**Figure 2:**
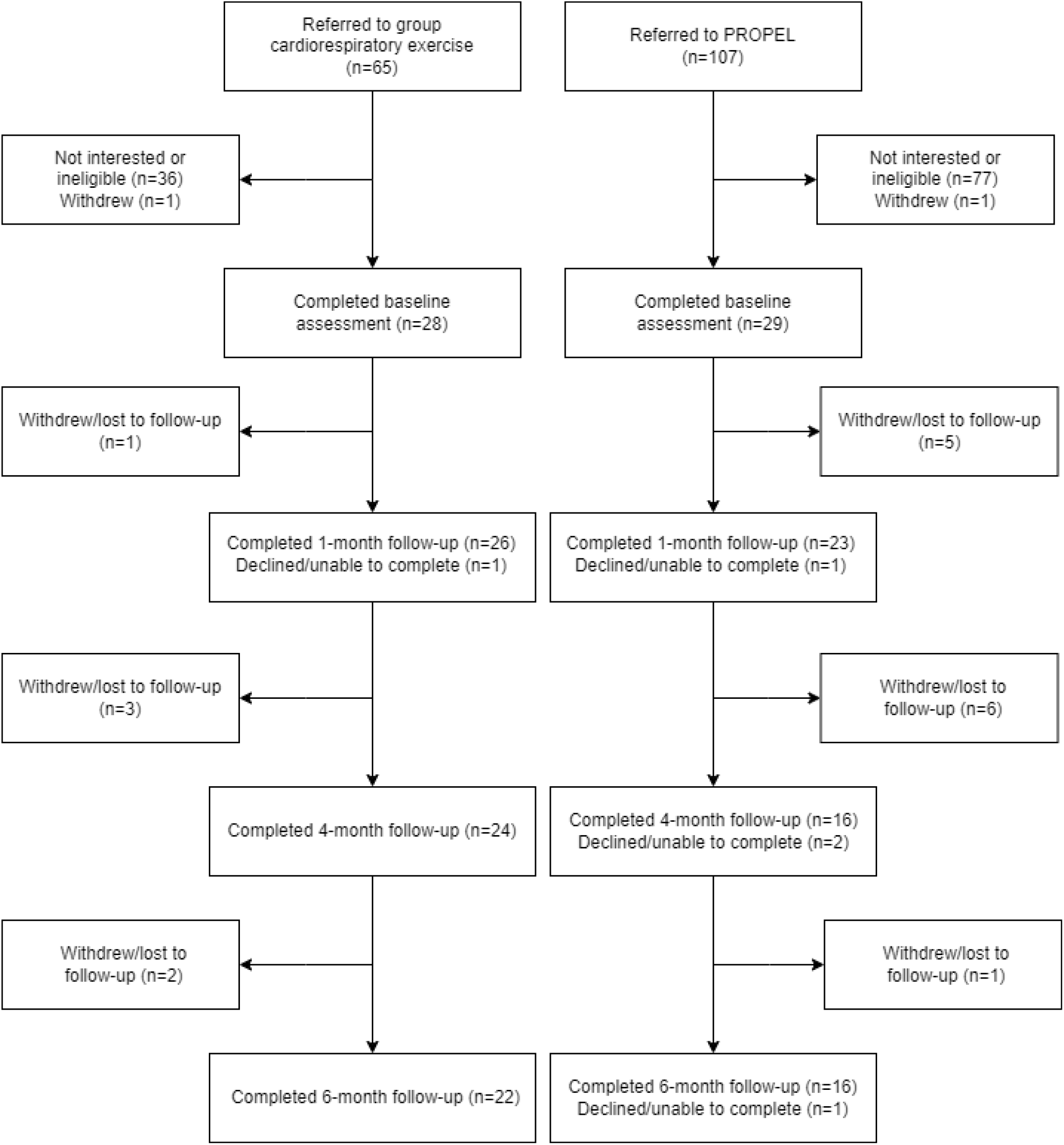
Participant flow after enrolment.

### Physical activity and exercise

At each time point, 27-42% of participants met the guidelines for physical activity (Table 2). There was no statistically significant difference between phases (GCE and PROPEL) in the odds of meeting physical activity guidelines (p-values>0.84; Table 3).

**Table 2:** Adherence to physical activity guidelines post-discharge. Values presented are the proportion of participants who met the guidelines for physical activity at each time point for each group. Note that missing data were imputed for participants who declined or were unable to complete the assessment at each time point, but had not yet withdrawn from the study.

|  | GCE | PROPEL |
| --- | --- | --- |
| 1 month | 7/27 | 10/24 |
| 4 months | 11/24 | 8/18 |
| 6 months | 9/22 | 6/17 |

GCE: Group Cardiorespiratory Exercise; PROPEL: Promoting Optimal Physical Exercise for Life

**Table 3:** Odds of meeting physical activity guidelines.

|  | Odds ratio | p-value |
| --- | --- | --- |
| Time (months) | 1.1 [0.92, 1.30] | 0.32 |
| Phase |  |  |
| GCE | Reference |  |
| PROPEL | 1.16 [0.29, 4.70] | 0.84 |
| Site |  |  |
| A | 0.91 [0.16, 5.15] | 0.95 |
| B | 1.15 [0.26, 5.02] | 0.86 |
| C | 3.08 [0.81, 11.78] | 0.10 |
| D | 0.62 [0.18, 2.19] | 0.46 |
| F | Reference |  |

**Table 4:** Physical activity 1, 4, and 6 months post-discharge intervention. Values presented are means with standard deviation in parentheses. The p-values are from analysis of covariance, with site included as a covariate.

|  | 1 month |  | 4 months |  | 6 months |  | *p-values* |  |
| --- | --- | --- | --- | --- | --- | --- | --- | --- |
|  | GCE<br>(n=27) | PROPEL<br>(n=24) | GCE<br>(n=24) | PROPEL<br>(n=18) | GCE<br>(n=22) | PROPEL<br>(n=17) | *Phase* | *Time* |
| Walking activity<br>(steps/day) | 4725<br>(5970) | 5825<br>(6470) | 5633<br>(5930) | 5643<br>(5332) | 5156<br>(5277) | 6728<br>(8150) | 0.97 | 0.35 |
| Time in<br>moderate-vigorous heart<br>rate range<br>(minutes/day) | 26.1<br>(36.3) | 94.1<br>(300.4) | 34.6<br>(58.2) | 29.6<br>(39.9) | 29.3<br>(39.5) | 40.5<br>(73.6) | 0.034 | 0.59 |
| Time in vigorous<br>heart rate range<br>(minutes/day) | 5.0 (9.8) | 22.6<br>(82.6) | 8.7 (15.9) | 5.5 (7.6) | 7.3 (13.9) | 6.5 (11.8) | 0.011 | 0.60 |
| PASIPD (MET<br>hours/day) | 12.0<br>(19.5) | 8.6 (9.3) | 8.9 (8.1) | 9.6 (7.3) | 7.9 (5.8) | 9.2 (7.0) | 0.60 | 0.46 |

GCE: Group Cardiorespiratory Exercise; MET: metabolic equivalent for time; PASIPD: Physical Activity Scale for Individuals with Physical Disabilities; PROPEL: Promoting Optimal Physical Exercise for Life.

#### Self-efficacy for exercise, outcome expectations, and barriers to exercise

Participants with stroke who completed PROPEL reported higher self-efficacy (p=0.0095), but not outcome expectations for exercise (p=0.92). There was no statistically significant difference between phases for the total number of reported barriers to exercise participation (p = 0.86), or for any of the individual components of the BBAQ (Table 5).

**Table 5:**
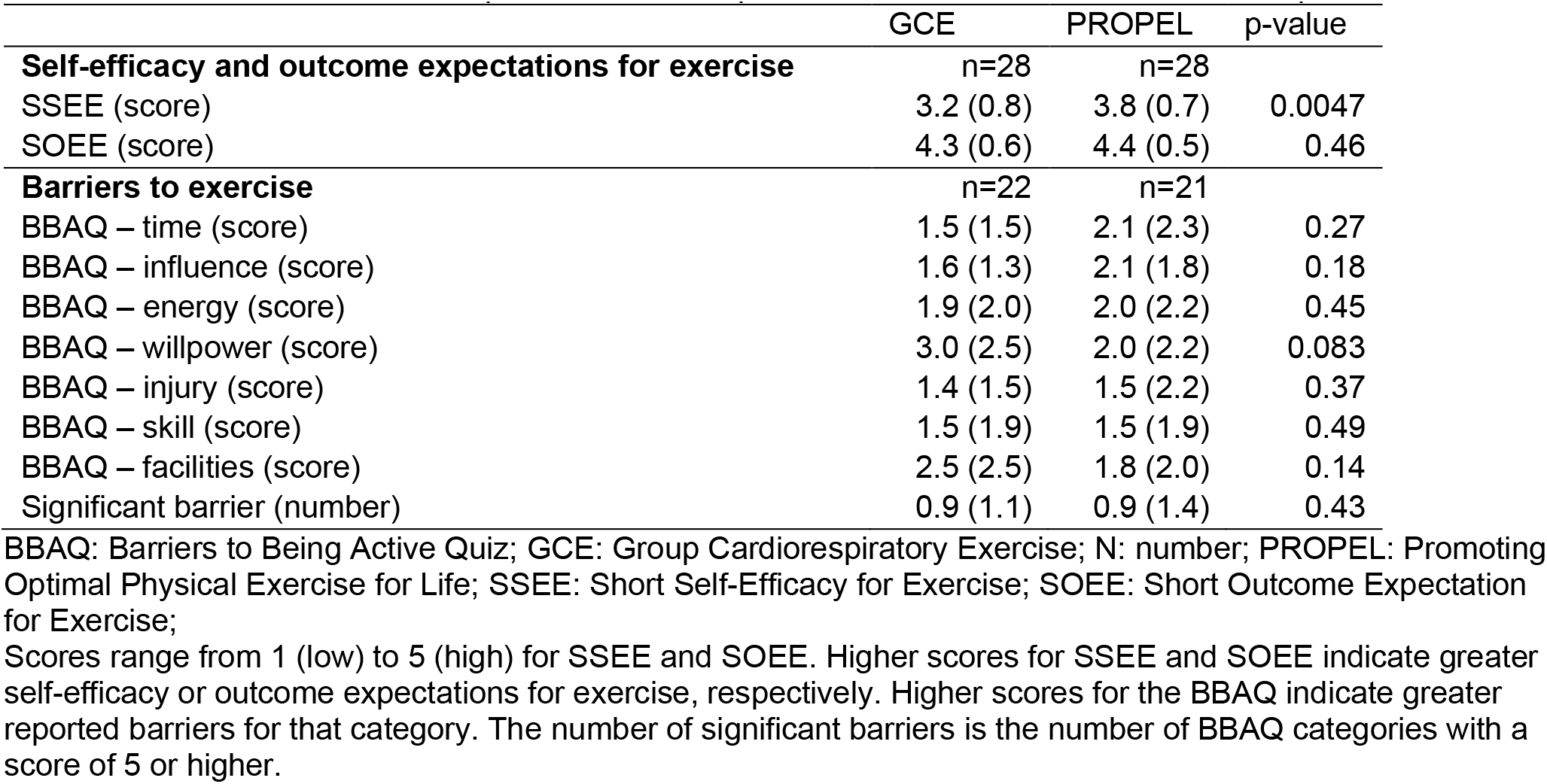
Self-efficacy for exercise, outcome expectations, and barriers to exercise. Values presented are means with standard deviations in parentheses. The p-values are from the Wilcoxon two-sample tests.

|  | GCE | PROPEL | p-value |
| --- | --- | --- | --- |
| **Self-efficacy and outcome expectations for exercise** | n=28 | n=28 |  |
| SSEE (score) | 3.2 (0.8) | 3.8 (0.7) | 0.0047 |
| SOEE (score) | 4.3 (0.6) | 4.4 (0.5) | 0.46 |
| **Barriers to exercise** | n=22 | n=21 |  |
| BBAQ – time (score) | 1.5 (1.5) | 2.1 (2.3) | 0.27 |
| BBAQ – influence (score) | 1.6 (1.3) | 2.1 (1.8) | 0.18 |
| BBAQ – energy (score) | 1.9 (2.0) | 2.0 (2.2) | 0.45 |
| BBAQ – willpower (score) | 3.0 (2.5) | 2.0 (2.2) | 0.083 |
| BBAQ – injury (score) | 1.4 (1.5) | 1.5 (2.2) | 0.37 |
| BBAQ – skill (score) | 1.5 (1.9) | 1.5 (1.9) | 0.49 |
| BBAQ – facilities (score) | 2.5 (2.5) | 1.8 (2.0) | 0.14 |
| Significant barrier (number) | 0.9 (1.1) | 0.9 (1.4) | 0.43 |

BBAQ: Barriers to Being Active Quiz; GCE: Group Cardiorespiratory Exercise; N: number; PROPEL: Promoting Optimal Physical Exercise for Life; SSEE: Short Self-Efficacy for Exercise; SOEE: Short Outcome Expectation for Exercise;
Scores range from 1 (low) to 5 (high) for SSEE and SOEE. Higher scores for SSEE and SOEE indicate greater self-efficacy or outcome expectations for exercise, respectively. Higher scores for the BBAQ indicate greater reported barriers for that category. The number of significant barriers is the number of BBAQ categories with a score of 5 or higher.

### Harms

There were no intervention-related adverse events reported during either GCE or PROPEL phases.

## DISCUSSION

This step-wedge randomized controlled trial aimed to determine whether a physical activity behaviour change intervention (PROPEL) leads to increased participation in physical activity and exercise in the community 6 months after discharge from rehabilitation post-stroke, compared to those who only participated in cardiorespiratory exercise in a group setting (GCE). We found that 27-42% of participants met the recommended guidelines for physical activity at each of the 1, 4, or 6-month time points post-stroke. However, there was no statistically significant difference between PROPEL and GCE participants in the odds of meeting physical activity guidelines. While people with stroke who completed PROPEL spent more time than GCE participants in the moderate-vigorous and vigorous heart-rate ranges post-discharge from rehabilitation, there was no difference between phases in walking activity or self-reported physical activity. We also aimed to determine whether those who completed PROPEL had higher self-efficacy for exercise, greater expectations of positive outcomes from exercise, and report fewer barriers to participating in exercise. People with stroke who completed PROPEL reported higher self-efficacy, but not outcome expectations, for exercise. There were no differences in the number of barriers to exercise participation, in addition to the specific characteristics of barriers to being physically active, including active time, influence, energy, willpower, injury, skill, and facilities, between the two groups.

To maximize the reliability of our findings, we used data from the activity monitor (step count and heart rate) and physical activity questionnaire to obtain an objective measure of post-discharge physical activity adherence for people with stroke. We used the standard within-cluster multiple imputation strategy (42) to handle missing data for participants who were still enrolled in the study at each time point. We found that participants who completed PROPEL spent more time per day in the moderate-vigorous and vigorous heart rate ranges post-discharge than those who completed GCE alone. While the difference between phases was small at some time points (e.g., PROPEL participants spent 10 mins/day more than GCE participants in moderate-vigorous heart rate range), these small increases in physical activity may still be meaningful. A meta-analysis of individual level physical activity data found a dose-response association between increased levels of physical activity and decreased risk of premature mortality in middle aged and older adults, with an increase in moderate-vigorous activity of 10 mins/day associated with a 44% decrease in all-cause mortality (43).

PROPEL was a supervised and individualized cardiorespiratory training that consisted of a behaviour change program, developed by integrating principles from the Transtheoretical Model (44) and Social Cognitive Theory (45), with an aim to increase exercise self-efficacy (20) and participation in exercise. The SSEE measures an individual’s confidence in their ability to exercise through pain, fatigue, being alone, and feeling depressed (34, 46, 47). A higher score indicates a greater self-efficacy for exercise. In our study, people with stroke who completed PROPEL reported higher self-efficacy for exercise when compared to those who completed GCE only (when measured at the time of enrolment i.e., after completing GCE or PROPEL). However, higher self-efficacy for exercise did not translate into improvements in the objective or self-reported measurements of physical activity post-discharge from rehabilitation in the PROPEL phase. Previously, Resnick and Jenkins (48) reported that higher self-efficacy for exercise measured using the SSEE was associated with increased exercise activity among older adults. In stroke survivors, self-efficacy for exercise reported through the SSEE was significantly associated with exercise behaviour, which accounted for 13% of the variance in exercise participation (34). It is possible that the objective measures used in our study did not capture the increased physical activity levels due to technological challenges, such as the inability to record the minutes of activity through Fitbit. Alternatively, there may be a need to further investigate the complex nature of exercise behaviour post-stroke through theoretical mediators of exercise adherence other than self-efficacy and outcome expectations (49). For instance, the SSEE asks more about ‘intrinsic’ barriers to exercise such as pain and fatigue. Perhaps people with stroke still experience external barriers, such as lack of access to facilities, as evident through the BBAQ scores (Table 5). Future trials should focus on addressing these barriers to translate the improvements in self-efficacy for exercise into increases in physical activity in the long term.

Participants with stroke in PROPEL phase reported a higher self-efficacy for exercise at discharge and spent more time in the moderate-to-vigorous and vigorous heart rate ranges 6-months post-discharge from rehabilitation than participants enrolled during the GCE phase. However, there were no differences between the phases for steps taken per day or duration of physical activity post-discharge. Factors at discharge after stroke rehabilitation such as age, pre-stroke activity, ambulation self-efficacy, perceived recovery from stroke, perceived health outcomes, walking speed and endurance, fatigue, mood, and executive function, have independently predicted walking activity in the community after stroke (9). Among these factors, Mahendran, Kuys (9) et al. suggested that walking endurance should be addressed during inpatient stroke rehabilitation, as higher walking endurance at discharge is linked to increased walking activity in the first month after discharge. The complex interaction between factors such as fatigue and walking activity post-stroke highlights the need for the integration of social/behavioral and self-efficacy components into the physical activity interventions post-stroke (50). To this end, there is a need to develop novel physical interventions based on theoretical frameworks that are relevant to increasing the duration of self-directed physical activity after discharge from inpatient stroke rehabilitation through goal-setting, problem-solving, and seeking support (51).

An important limitation of this study is that few participants were enrolled in the interventions, GCE and PROPEL. Approximately 3,000 people with stroke were admitted across all sited combined during the study. However, only 172 patients (i.e., ∼6% of admissions) were referred to the GCE or PROPEL programs. This is much lower than the expected ∼40% of patients who should be eligible for cardiorespiratory exercise early post-stroke (6). Because of the lower rate of referral to the programs, the research PTs reported a challenge to get at least three participants to start a group; therefore, some eligible patients referred to the programs may have been lost as a group could not be formed before they were discharged from rehabilitation. Due to the combined effect of lower rate of referral to the exercise groups, requirement to have a closed group of three participants, and early termination of the study due to the COVID-19 pandemic, only 25% of the target sample size was recruited for this study. However, based on the trends observed from this small sample (e.g., Table 2), it is unlikely that group differences would have been observed with a larger sample size. The study sample predominantly consisted, on average, of participants with mild post-stroke impairment (e.g., NIH-SS ∼ 3, CMSA ∼ 5), limiting our ability to generalize the study findings to the people with more severe strokes.

In a stepped wedge study design, blinding research personnel to intervention allocation may be compromised as it is more likely that a site is delivering one type of intervention at a time as the trial progresses (52). This situation could increase the risk of unconscious selection bias, even though the clusters were randomly allocated over time. For example, the PTs who referred participants to the program may have been more likely to refer those with challenges to exercise adherence to the program if they believed that the site was running the PROPEL program at that time, as they were aware that PROPEL had an additional educational component to improve adherence. Finally, factors such as seasonal variation in physical activity levels, post-pandemic reduction in physical activity, and differences in geographical barriers to physical activity between study locations, could have influenced the results.

## Conclusions

Participation in the PROPEL program during rehabilitation after stroke increases self-efficacy for exercise immediately after completing PROPEL when compared to participation in GCE alone. Completing the PROPEL program may not lead to patients with stroke meeting the recommended duration and intensity of physical activity. However, patients with stroke who completed PROPEL spent more time per day in the moderate-vigorous and vigorous heart rate ranges until six months post-discharge than those who completed GCE alone.

## Data Availability

The participants of this study did not give consent for their data to be shared publicly, so supporting/raw data is not available.

## Acknowledgements

We thank Michelle Legasto, Rina Reyes, Julie Lo, and Jayne Hall, who delivered the intervention at the study sites, and Kay-Ann Allen, Anthony Aqui, and Laura Fabiano, who recruited study participants and collected the data. We thank Louis Biasin and Vivien Poon who contributed to the development of the Promoting Optimal Physical Exercise for Life (PROPEL) program.

## SUPPLEMENTARY MATERIAL: COMPLETE CASE ANALYSIS

**Supplementary Table 1:**
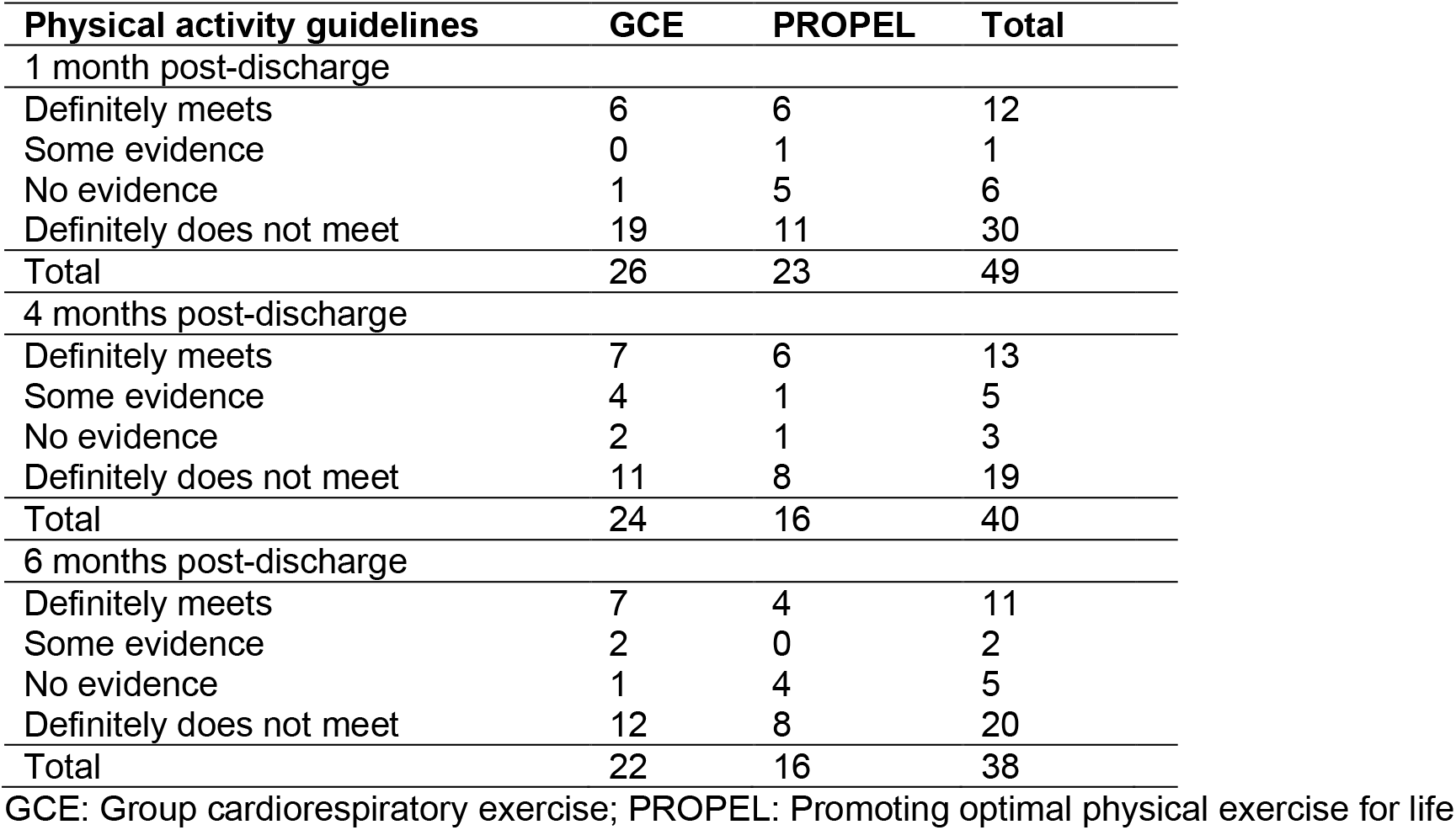
Frequency of meeting physical activity guidelines between groups – complete case analysis. Values presented are the number of participants meeting the guidelines at each time point. Participants were deemed to meet the recommended intensity and duration of physical activity in a given week if they met the physical activity guidelines according to at least 2 of the three modalities (step count, heart rate, and PASIPD data, that is, at least 6000 steps per day, or at least 150 minutes per week of moderate to vigorous intensity exercise, or at least 75 minutes per week of vigorous-intensity exercise). At each time point, participants were classified as (a) definitely meets guidelines (meets guidelines in 2/3 or 2/2 available modalities); (b) some evidence for meeting guidelines (meets guidelines in 1/2 or 1/1 available modalities); (c) no evidence for meeting guidelines (meets guidelines in 0/1 modalities), or (d) definitely does not meet guidelines (meets guidelines in ≤1/3 or 0/2 available modalities).

| Physical activity guidelines | GCE | PROPEL | Total |
| --- | --- | --- | --- |
| 1 month post-discharge |  |  |  |
| Definitely meets | 6 | 6 | 12 |
| Some evidence | 0 | 1 | 1 |
| No evidence | 1 | 5 | 6 |
| Definitely does not meet | 19 | 11 | 30 |
| Total | 26 | 23 | 49 |
| 4 months post-discharge |  |  |  |
| Definitely meets | 7 | 6 | 13 |
| Some evidence | 4 | 1 | 5 |
| No evidence | 2 | 1 | 3 |
| Definitely does not meet | 11 | 8 | 19 |
| Total | 24 | 16 | 40 |
| 6 months post-discharge |  |  |  |
| Definitely meets | 7 | 4 | 11 |
| Some evidence | 2 | 0 | 2 |
| No evidence | 1 | 4 | 5 |
| Definitely does not meet | 12 | 8 | 20 |
| Total | 22 | 16 | 38 |

GCE: Group cardiorespiratory exercise; PROPEL: Promoting optimal physical exercise for life

**Supplementary Table 2:** Odds of meeting physical activity guidelines. The analysis only included participants who definitely met or definitely did not meet the physical activity guidelines, as defined in Supplementary Table 2.

|  | Odds ratio | p-value |
| --- | --- | --- |
| Time (months) | 1.10 [0.91, 1.30] | 0.31 |
| Phase |  |  |
| GCE | Reference |  |
| PROPEL | 0.37 [0.04, 3.67] | 0.40 |
| Site |  |  |
| A | 0.37 [0.76, 2.25] | 0.28 |
| B | 1.45 [0.21, 9.87] | 0.70 |
| C | 2.18 [0.37, 12.76] | 0.39 |
| D | 3.73 [0.18, 41.65] | 0.40 |
| F | Reference |  |

**Supplementary Table 3:** Comparison of physical activity data between groups – complete case analysis. Values presented are means with standard deviations (SD) in parentheses.

|  | 1 month |  | 4 months |  | 6 months |  | *p-values* |  |
| --- | --- | --- | --- | --- | --- | --- | --- | --- |
|  | GCE<br>(n=26)* | PROPEL<br>(n=23)<sup>†</sup> | GCE<br>(n=24)<sup>‡</sup> | PROPEL<br>(n=16)<sup>§</sup> | GCE<br>(n=22)** | PROPEL<br>(n=16)<sup>††</sup> | *Phase* | *Time* |
| Walking activity<br>(steps/day) | 4138<br>(4522) | 4253<br>(3028) | 5033<br>(4002) | 5168<br>(3179) | 4420<br>(3046) | 4720<br>(3493) | 0.080 | 0.19 |
| Time in<br>moderate-vigorous heart<br>rate range<br>(minutes/day) | 23.3<br>(29.5) | 27.8<br>(33.0) | 28.8<br>(45.0) | 29.3<br>(24.8) | 25.2<br>(29.0) | 33.5<br>(51.0) | 0.64 | 0.37 |
| Time in vigorous<br>heart rate range<br>(minutes/day) | 3.7 (6.0) | 7.1 (14.2) | 7.1 (12.6) | 4.8 (4.9) | 5.7 (9.2) | 5.2 (8.2) | 0.23 | 0.52 |
| PASIPD (MET<br>hours/day) | 8.3 (6.1) | 8.3 (7.2) | 9.3 (7.6) | 10.5 (5.2) | 8.1 (4.9) | 9.0 (5.4) | 0.71 | 0.94 |

\* n=25 for Fitbit data (steps/day and time in heart rate ranges); n=22 for PASIPD scores
<sup>†</sup> n=21 for PASIPD scores and 18 for Fitbit data<sup>‡</sup> n=21 for PASIPD scores, n=22 for steps/day, and n=20 for time in heart rate ranges<sup>§</sup> n=14 for Fitbit data<sup>\*\*</sup> 22 participants were included in the analysis, but data were only available for 20 participants for each variable<sup>††</sup> n=12 for Fitbit data

## Notes

**Funding:** This study was supported by the Canadian Institutes of Health Research (PJT148906). AM held a New Investigator Award from the Canadian Institutes of Health Research (MSH141983). AT was supported by a personnel award from the Heart and Stroke Foundation, Ontario Provincial Office (CS I 7468). The funding source had no role in the design or execution of the study, analyses, or interpretation of the data or decision to submit results.

**Declaration of interest:** The authors report no conflicts of interest.

### Competing Interest Statement

The authors have declared no competing interest.

### Clinical Trial

NCT02951338

### Clinical Protocols

https://bmjopen.bmj.com/content/7/6/e015843

### Funding Statement

This study was supported by the Canadian Institutes of Health Research (PJT148906). AM held a New Investigator Award from the Canadian Institutes of Health Research (MSH141983). AT was supported by a personnel award from the Heart and Stroke Foundation, Ontario Provincial Office (CS I 7468). The funding source had no role in the design or execution of the study, analyses, or interpretation of the data or decision to submit results.

### Author Declarations

This study was approved by the research ethics boards of: (a) Hamilton Health Sciences, Hamilton, Ontario (approval number: 2274); (b) Sunnybrook Health Sciences Centre, Toronto, Ontario (approval number: 472-2016); (c) St. Joseph's Care Group, Thunder Bay, Ontario (approval number: 2016011); (d) University Health Network, Toronto, Ontario (approval number: 16-5916); (e) West Park Healthcare Centre, Toronto, Ontario (approval number not provided in the letter); (f) Thunder Bay Regional Health Sciences Centre, Thunder Bay, Ontario (approval number: 2016139).

